# Towards repeatable and converging methods in diffusion MRI: Evidence from a longitudinal chronic pain cohort

**DOI:** 10.1101/2025.10.03.25337290

**Authors:** Marc-Antoine Fortier, Graham Little, Paul Bautin, Monica Sean, Pascal Tétreault

## Abstract

**Introduction:** Studies of white matter (WM) alterations in chronic low back pain (CLBP) using diffusion MRI (dMRI) have yielded inconsistent results, highlighting a need for more reproducible methods. This study introduces a longitudinal analysis pipeline designed to identify stable and repeatable WM microstructural differences between CLBP patients and healthy controls.

**Methods:** Diffusion MRI was acquired from 27 CLBP patients and 25 control participants at three separate visits with two-month intervals. A customized Tract-Based Spatial Statistics (TBSS) analysis was performed on fractional anisotropy (FA) maps that were averaged across visits for each participant. Resulting clusters showing group differences were subsequently filtered based on cluster size (>10 voxels) and a high repeatability threshold (image intra-class correlation coefficient [I2C2] > 0.75) to ensure findings were stable across all three imaging sessions. Results were compared against standard single-visit analyses and an alternative tractometry analysis.

**Results:** The repeatability analysis identified 11 clusters with stable and significant FA differences. Seven clusters, located primarily in the occipital, parietal, and frontal lobes, showed higher FA in controls. Four clusters, located in the frontal and temporal lobes, showed higher FA in the CLBP group. In contrast, single-visit analyses identified a much larger number of clusters (21 to 36), the majority of which were not spatially consistent across time and did not overlap with the final repeatable clusters. The repeatable TBSS findings demonstrated a strong spatial correspondence with group differences found using a tractometry analysis.

**Discussion:** By incorporating a longitudinal design and explicit repeatability filtering, our method effectively reduces spurious findings common in single-visit dMRI studies. This approach successfully isolated reliable WM regions with altered dMRI metrics in CLBP, demonstrating its value in improving the robustness of neuroimaging research in chronic pain.

## 1 Introduction

The association of chronic pain (CP) to alterations in human brain structure and function has become a well established fact (Baliki & Apkarian, 2015). In-vivo magnetic resonance imaging studies have demonstrated CP related alterations to brain function and structure but have primarily focused on human grey matter (Kregel et al., 2015; van der Miesen et al., 2019) . As understanding of CP has evolved, CP has been theorized to be more broadly considered a network disorder, warranting the study of the human brain’s white matter which encompasses the microstructural fibers that transfer information between disparate regions of the brain. Diffusion magnetic resonance imaging (dMRI) is a technique that measures the anisotropic movement of water molecules in-vivo and is sensitive to subtle changes to white matter microstructure (Beaulieu, 2002). Historically in the CP literature, the most common approach of inferring microstructural alterations in human white matter (WM) with dMRI data, is to fit a three dimensional tensor and report parametric values extracted from the tensor such as fractional anisotropy (FA). Fractional anisotropy is a metric that reflects the variation of diffusion along the tensor’s primary axis relative to the perpendicular axes, and is related to the underlying microstructural organization/composition of tissue within a voxel. To date, studies using dMRI to examine human WM as it relates to chronic pain remain scarce and have produced inconclusive results (Bautin et al., 2025). Results differ considerably between primary chronic pain conditions (e.g. chronic primary headache pain, chronic primary visceral pain, chronic primary musculoskeletal pain) with a multitude of dMRI acquisition and analysis methods implemented, leading to difficulty in comparing and interpreting differences between studies.

Even in studies where specific pain conditions were targeted (e.g. chronic low back pain (CLBP) and subacute back pain (SBP)), various analysis methodology were used and notable inconsistencies have been reported in the location and the degree of WM FA group differences with a more consistent finding of lower FA in CLBP patients relative to controls. Three studies have shown reduced baseline FA in CLBP compared to controls with varying sample sizes; 102 CLBP patients, 50 controls (Kim et al., 2020); 24 CLBP patients, 22 controls (Ma et al., 2020); 31 CLBP patients, 31 controls (Mansour et al., 2013). Two studies have reported post-therapeutic effects measurable by dMRI in CLBP patients with FA increasing in WM regions after therapy (Čeko et al., 2015; Kim et al., 2020). While a study of patients with subacute back pain, successfully predicted those who transitioned to a chronic pain state using measurements of FA along WM tracts (Mansour et al., 2013), with a followup study more robustly predicting this transition using dMRI tractography derived measures of connectivity (Vachon-Presseau et al., 2016). While some agreement is found among reports of lower FA associated with CLBP, regions implicated in CLBP vary substantially between studies; left insular WM (Čeko et al., 2015); primary somatosensory cortex white matter (Kim et al., 2020); corpus callosum, bilateral anterior thalamic radiation, right posterior thalamic radiation, right superior longitudinal fasciculus, and the left anterior corona radiata (Ma et al., 2020); the left superior longitudinal fasciculus, the left internal and external capsules, and anterior corona radiata (Mansour et al., 2013).

The problem of reproducible results is not confined to the field of chronic pain but is one of the fundamental challenges facing the neuroimaging field at large (Botvinik-Nezer & Wager, 2023). Many studies have addressed repeatability in test-retest studies of dMRI measurements reporting generally reliable dMRI measures in healthy adult controls (Ades-Aron et al., 2025; Boekel et al., 2017; Duan et al., 2015), across various age ranges (Luque Laguna et al., 2020; Merisaari et al., 2019; Rosberg et al., 2022) and in patient cohorts (Cole et al., 2014; Cousineau, Jodoin, Morency, et al., 2017; Keihaninejad et al., 2013) with FA showing the highest reliability among diffusion tensor measurements in most cases. Repeatability has also been studied longitudinally, demonstrating that more advanced dMRI modeling produces metrics that decrease in repeatability over a 3 month interval but repeatability remains at or above high levels (Newman et al., 2020). However, while these studies importantly assess the repeatability of dMRI metrics within controls and patients separately, to reconcile the inconsistent findings in the CP neuroimaging literature it is important to identify regional WM differences between controls and CP patients that can be found repeatedly at multiple time points.

Here we present a method of experimental design and voxel-wise analysis that uses the stability of dMRI derived measurements of white matter microstructure as a means to filter regional differences to only include group differences that are repeatable across three scans. The proposed repeatability analysis was applied to a pilot cohort of 25 controls and 27 CLBP patients with imaging acquired at 3 separate visits. As a comparison to the most commonly reported voxel-wise analysis method in the CP literature, we chose to use tract-based spatial statistics (TBSS), projecting adjacent FA values to a voxel-wise skeleton of a subject’s WM. Regions (clusters) that were found to have statistically significant between group differences were further filtered by image intra-class correlation coefficient (I2C2) values to remove regions that had non-repeatable measurements. In total, 11 regions were observed to have FA values that were stable across visits and were different between CLBP and control groups (7 higher in controls, 4 higher in CLBP). To compare our results to other more advanced methodology, an exploratory analysis was performed showing strong correspondence between the proposed repeatability analysis and tractometry, a method capable of segmenting and analyzing individual WM tracts from dMRI. The proposed repeatability analysis technique provides a method to incorporate reproducibility metrics into the analysis design and in the context of the CLBP pilot imaging study, detects regions with repeatable CLBP related group differences in FA, reducing the chance of spurious findings.

## 2 Methods

### 2.1 Participants

This study was approved by the ethics review board of the Centre intégré universitaire de santé et de services sociaux de l’Estrie – Centre hospitalier universitaire de Sherbrooke (CIUSSS de l’Estrie – CHUS), Sherbrooke, Canada (file number: 2021-3861). The study is also registered on the Open Science Framework (“Pilot project on brain and lower back imaging of chronic pain”, https://doi.org/10.17605/OSF.IO/P2Z6Y).

Control and CLBP participants recruited and asked to visit the research center of the CHUS for cerebral and lumbar MRI scans for a total of three visits with a two-month interval. Saliva samples and questionnaires were also collected as a part of the larger study. Apart from their usual treatment, no therapeutic intervention was modified or administered to participants between visits. All CLBP participants underwent pain intensity screening using the Brief Pain Inventory (BPI) short form (Poquet & Lin, 2016) to quantify their symptoms.

Exclusion and inclusion criteria for CLBP and HC with full details described in a previous study on grey matter density (Sean et al., 2024). In short, CLBP participants were included if they had low back pain for greater than 4 months, an average pain intensity of greater than or equal to 3/10 on the BPI within the 24-hour period before the initial visit and pain primarily localized to the low back. Control participants were excluded if they had a history of chronic pain, had pain at the time of testing or an outstanding painful episode within 3 months of enrollment.

In total, 27 CLBP (mean age 43 ± 16 years, 13 females) and 25 control participants (mean age 42 ± 14 years, 15 females) were recruited for this study, and had adequate imaging data for at least one of the 3 visits. Amongst these cohorts, 23 CLBP participants and 24 control participants completed all three visits or had adequate diffusion and anatomical imaging data acquired for the subsequent analysis.

### 2.2 MRI acquisition

Imaging data was acquired on a Philips Ingenia 3.0T MRI located at the CHUS. For each subject and visit, whole brain T1-weighted structural imaging and diffusion weighted imaging (DWI) were acquired. Structural T1-weighted imaging (1.0 × 1.0 × 1.0 mm³) was acquired with TE = 3.5 ms, TR = 7.9 ms, flip angle = 8°, and a scan time of 4:19 min. DWI was acquired with a single-shot EPI spin-echo sequence (Multiband factor = 2, SENSE reduction factor = 1.9, Partial Fourier (Halfscan) factor = 0.6949152), TR = 4800 ms, TE = 92 ms, flip angle = 90°, 66 slices were acquired at a 2 mm thickness with no gap and 2.0 × 2.0 mm² in-plane. In total, 108 diffusion volumes acquired using 7 b = 0 s/mm², 8 b = 300 s/mm², 32 b = 1000 s/mm², 60 b = 2000 s/mm², and 1 = 0 s/mm², acquired in the reversed phase encode direction in a scan time of 9:19 min.

### 2.3 Preprocessing

Structural T1 and diffusion weighted images for each subject and visit were preprocessed using the Tractoflow pipeline (Theaud et al., 2020). In short, the Tractoflow pipeline performs image denoising (Veraart et al., 2016), magnetic field susceptibility correction (Andersson et al., 2003), eddy current correction (Andersson & Sotiropoulos, 2016), registration from T1 imaging space to diffusion imaging space using ANTs (Avants et al., 2008), brain extraction and tissue segmentation on T1 images (Zhang et al., 2001), image intensity normalization (Tustison et al., 2010), spatial resolution resampling, outputting DTI parametric (e.g. fractional anisotropy (FA)) and fiber orientation distribution function (fODF) maps.

### 2.4 Tract-Based Spatial Statistics

To leverage the longitudinal design of the dataset, a customized repeatability analysis pipeline was used to extract regions (clusters) that had a measurable group difference at all three visits. The custom pipeline is visualized in Figure 1. Using the preprocessed FA maps output from the previous steps, Tract-Based Spatial Statistics (TBSS) (Smith et al., 2006), an automated voxelwise diffusion analysis tool (FMRIB software suite), was performed using default parameters by first generating a white matter skeleton, at a spatial resolution of 1 mm × 1 mm × 1 mm, for the entire cohort (all subjects and scans included) and then projecting FA values to this skeleton for each subject and scan.

**Figure 1.**
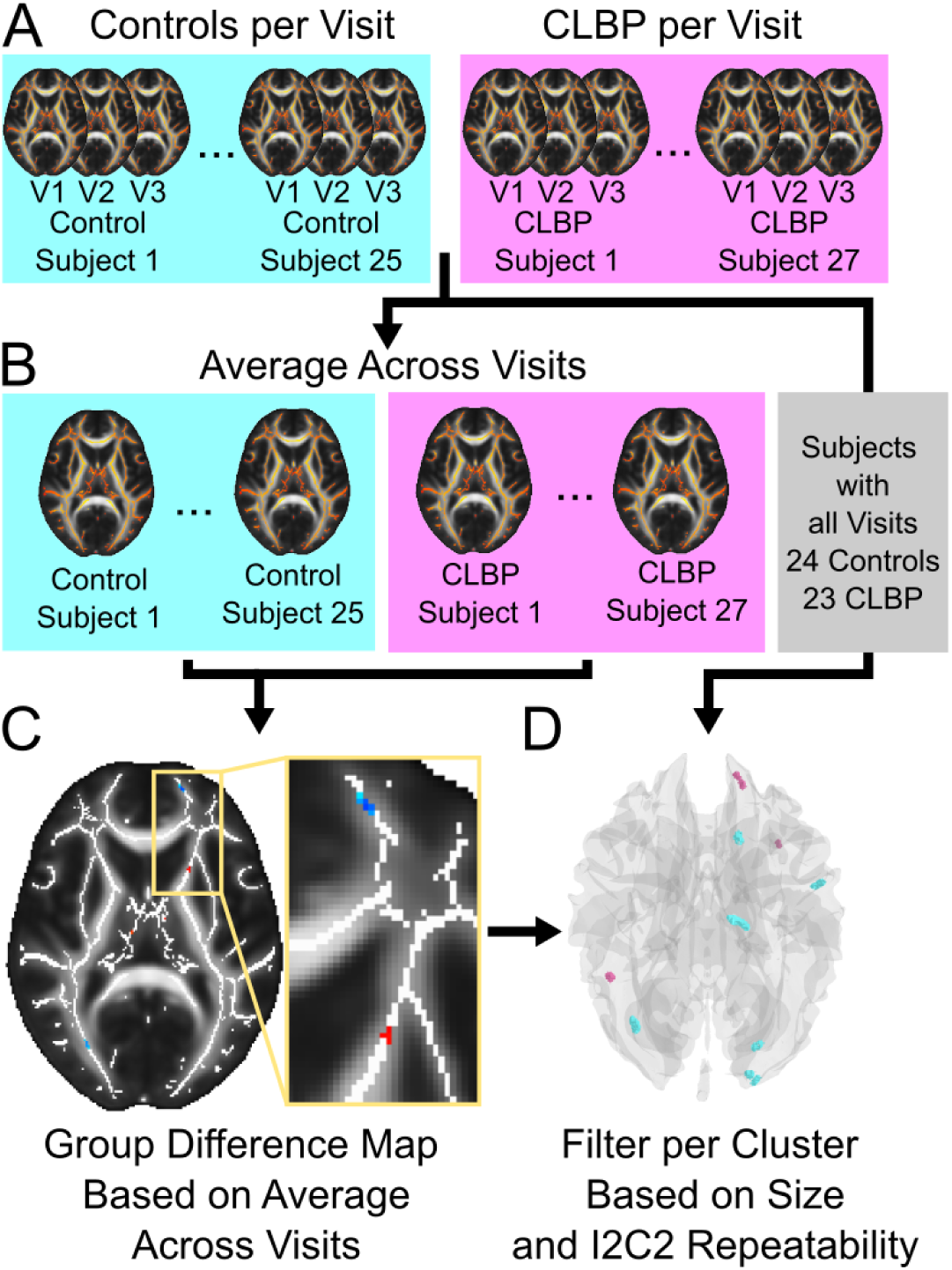
Workflow of analysis used to generate clusters with regional differences between controls and chronic low back pain (CLBP) subjects that are repeatedly observed across visits. A) 25 control subjects and 27 CLBP subjects were processed to generate skeletonized FA maps per subject for each visit (V1, V2, V2). B) Skeletonized FA maps were averaged across visits to attain a single map per subject. C) A group difference map was generated based on voxelwise t-tests and thresholded to include voxels with a t-statistic value greater than |2.5| (p < 0.0001). D) Clusters were filtered to include only clusters that were greater than 10 voxels in size and had an I2C2 value greater than 0.75. I2C2 values for each cluster were calculated using the subset of subjects that had imaging data available for all three visits.

### 2.5 Repeatability Analysis

To improve the sensitivity to detect group differences, skeletonized FA maps were first averaged across visits for each subject generating a single FA map per subject. A two-sample t-test was then performed (FSL randomise) between groups using the subject FA maps averaged across visits outputting a single parametric t-statistic map along the skeleton. The parametric map was thresholded at a t-statistic value of |2.5| (p <0.01) and remaining clusters were excluded if they were less than 10 voxels in size (i.e. a volume of 10 mm^3^, on 1mm^3^ white matter skeleton). Finally, to remove clusters that had unreliable FA values across visits, the image intra-class correlation (I2C2) (Shou et al., 2013) was calculated for each remaining cluster using only the subjects that had adequate imaging acquired at all 3 visits. Clusters with I2C2 values less than 0.75 (good to excellent reliability, (Koo & Li, 2016; Shou et al., 2013)) were removed leaving clusters with a measurable group difference and had repeatable values across visits.

### 2.6 Comparison to single visits

As a comparison to standard cross-sectional designs, the same group difference analysis, parameter map thresholding (t-statistic greater than |2.5|) and cluster size thresholding (clusters containing greater than 10 voxels) was performed separately for each visit. Resulting clusters were then separated into those where controls had greater FA compared to the CLBP group (t-statistic greater than 2.5) and those where CLBP had greater FA compared to the control group (t-statistic less than 2.5). Clusters retained from the three analyses performed for each visit separately were compared to those retained from the proposed repeatability analysis as follows. First, the total number of clusters retained after statistical/voxel size thresholding at each visit were totalled. Then the clusters retained at each visit were compared to those retained after the proposed repeatability analysis to identify the number of clusters that spatially overlap in both analyses. For example, for the visit 1 analysis, both the total number of clusters retained and the total number of clusters that overlap the proposed repeatability analysis are calculated.

### 2.7 Comparison to Alternative Methodology - Tractometry Analysis

To compare results from the proposed repeatability analysis with alternative methodology, a supplementary automated tractometry analysis was performed. Whole brain tractograms were generated using the previously used Tractoflow pipeline (Theaud et al., 2020) with default analysis parameters. Whole brain tractograms were then segmented into separate bundles using BundleSeg (St-Onge et al., 2023) with a whole brain white matter tract atlas (Rheault, 2023), outputting 51 white matter tracts per subject. Six clusters (4 control FA > CLBP FA in the occipital lobe and 2 CLBP FA > control FA located in the superior frontal lobe) were selected for further investigation from the TBSS repeatability analysis because they were located in adjacent regions. White matter tracts overlapping the selected clusters were retained for tractometry analysis. Each retained tract was segmented into 50 parcellations per subject per visit and average FA was calculated for each segment (Cousineau, Jodoin, Garyfallidis, et al., 2017). To test for segments with different FA values between controls and CLBP, two-sample t-tests (p < 0.05, uncorrected) were performed per segment per visit. For comparison to the repeatability analysis, tract segments that overlapped clusters output from the proposed analysis were identified.

## 3 Results

### 3.1 Group Average Regional Differences in Fractional Anisotropy

The repeatability analysis pipeline identified eleven white matter clusters with stable group differences in fractional anisotropy (FA) across all three visits. The eleven clusters are visualized in Figure 2, along with average FA values extracted for each cluster. Seven of these clusters showed significantly higher FA in the control group compared to the CLBP group, while four clusters showed higher FA in the CLBP group. The distribution of these FA values for each participant, averaged across the three visits, is plotted in Figure 2B (Controls > CLBP) and Figure 2C (CLBP > Controls), illustrating a clear separation between the groups.

**Figure 2.**
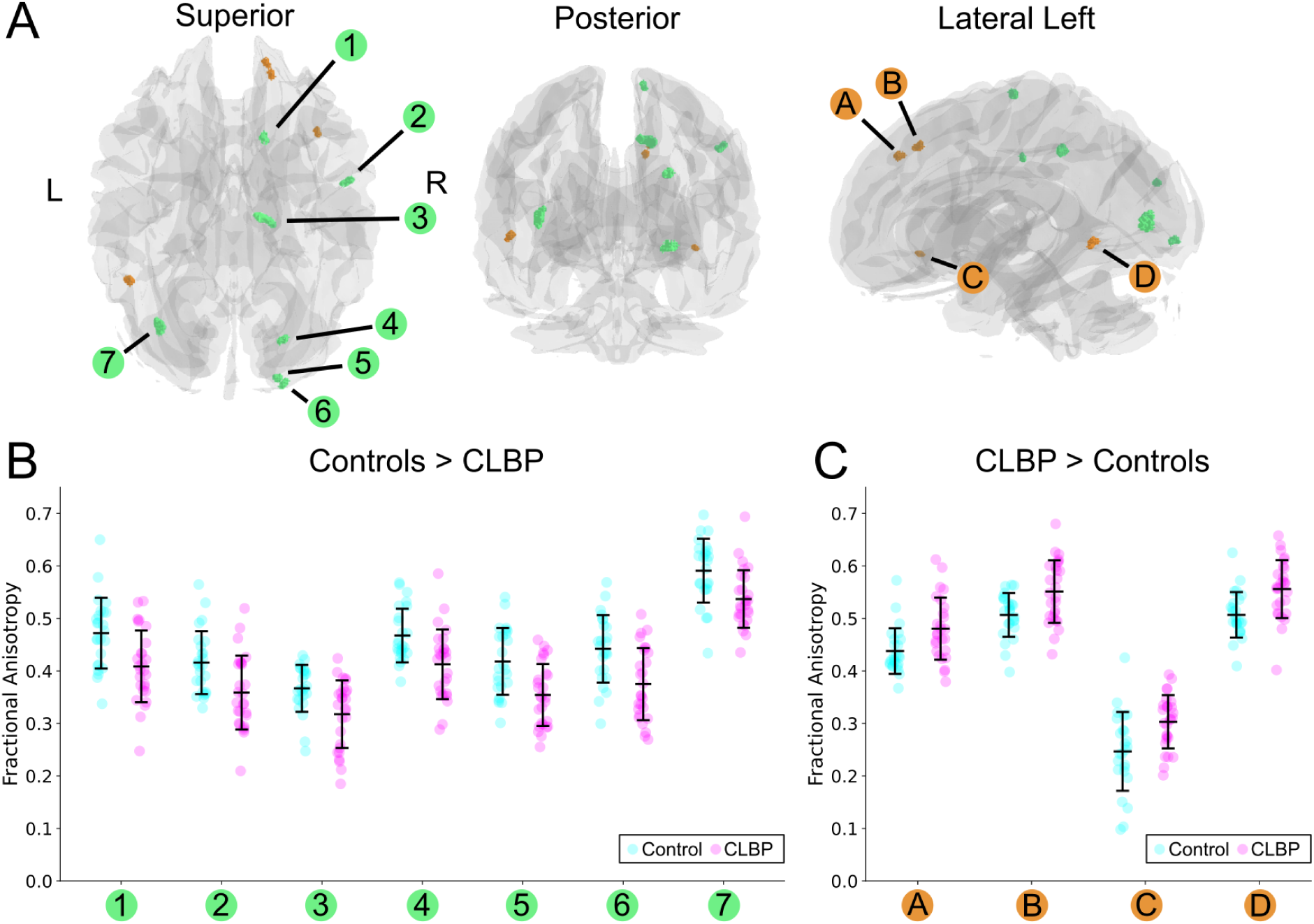
A) Visualization of clusters with different fractional anisotropy values between groups (t-statistic greater than |2.5|), cluster size of greater than 10 voxels and an I2C2 value greater than 0.75. Seven regions where fractional anisotropy was higher in controls relative to the chronic low back pain group are shown (green) along with 4 regions where fractional anisotropy was higher in the chronic low back pain group compared to controls (orange). Average fractional anisotropy values along with standard deviations (black) are shown for individual controls (cyan) and CLBP (pink) subjects averaged across visit for B) regions with higher fractional anisotropy in controls and C) regions with higher fractional anisotropy in the chronic low back pain group.

Among the 7 clusters showing higher FA in the control group relative to the control group, 4 were located in the occipital lobe (right hemisphere: clusters 4,5,6; left hemisphere; cluster 7), 2 were located in the right parietal lobe and 1 was located in the right frontal lobe. Among the 4 clusters showing higher FA in the CLBP group relative to controls, 3 were located in the right frontal lobe (clusters A, B, C) and cluster D located in the posterior portion of the left temporal lobe.

Detailed statistics for each of the eleven clusters are presented in Table 1. The clusters ranged in size from 10 mm³ to 48 mm³. Effect sizes were moderate to large, with Cohen’s D values ranging from 0.818 to 1.043 for clusters where controls had higher FA, and from -0.818 to -0.982 for clusters where CLBP patients had higher FA. All group differences were statistically significant (p < 0.005, uncorrected for multiple comparisons). All eleven clusters demonstrated good to excellent repeatability, with Image Intra-class Correlation Coefficient (I2C2) values for the entire cohort ranging from 0.7685 to 0.8821 (good to excellent (Koo & Li, 2016; Shou et al., 2013).)

**Table 1.**
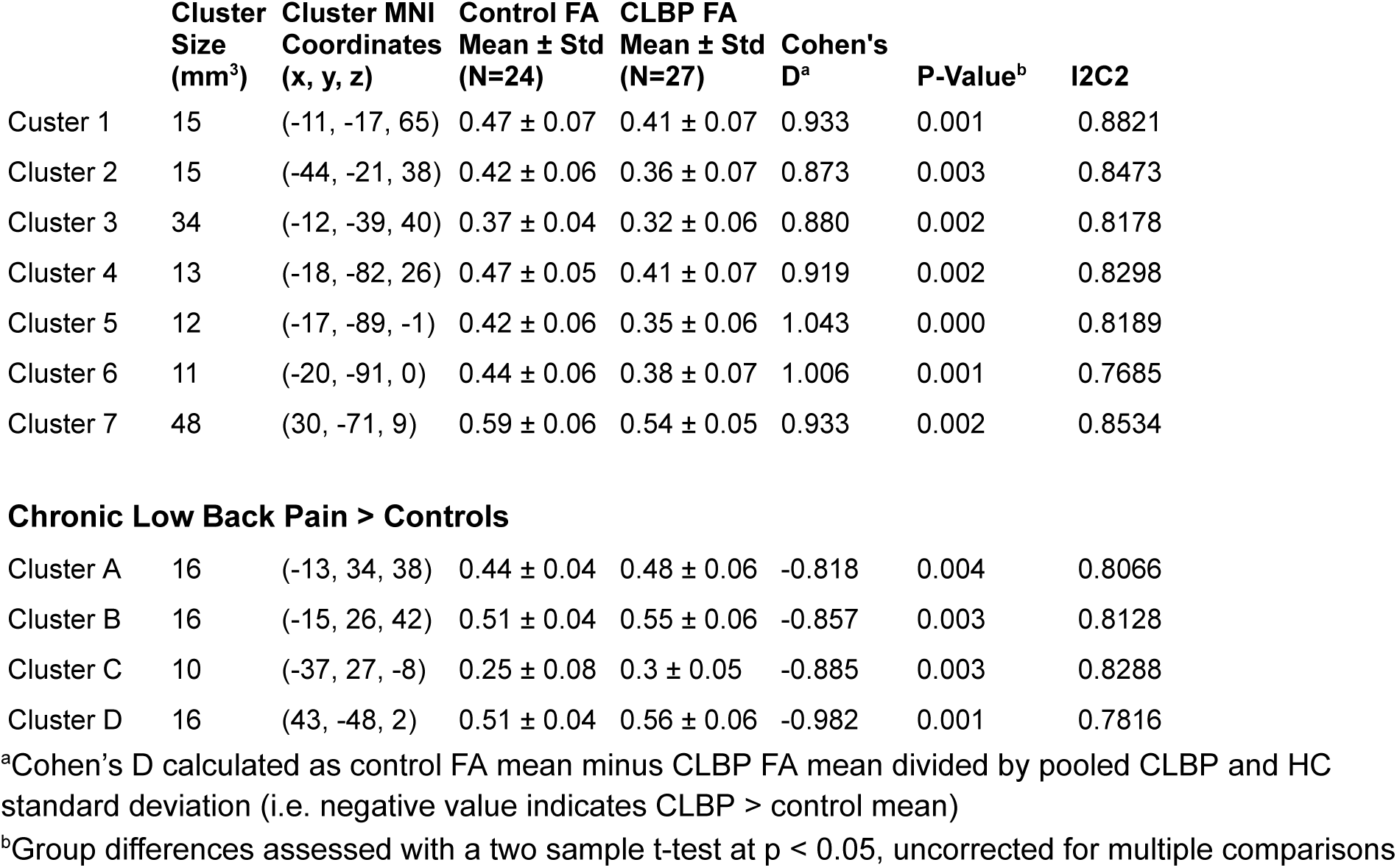
Group differences and repeatability of fractional anisotropy averaged for each cluster.

> Chronic Low Back Pain
|  | **Cluster Size (mm<sup>3</sup>)** | **Cluster MNI Coordinates (x, y, z)** | **Control FA Mean ± Std (N=24)** | **CLBP FA Mean ± Std (N=27)** | **Cohen's D<sup>a</sup>** | **P-Value<sup>b</sup>** | **I2C2** |
| --- | --- | --- | --- | --- | --- | --- | --- |
| Cluster 1 | 15 | (-11, -17, 65) | 0.47 ± 0.07 | 0.41 ± 0.07 | 0.933 | 0.001 | 0.8821 |
| Cluster 2 | 15 | (-44, -21, 38) | 0.42 ± 0.06 | 0.36 ± 0.07 | 0.873 | 0.003 | 0.8473 |
| Cluster 3 | 34 | (-12, -39, 40) | 0.37 ± 0.04 | 0.32 ± 0.06 | 0.880 | 0.002 | 0.8178 |
| Cluster 4 | 13 | (-18, -82, 26) | 0.47 ± 0.05 | 0.41 ± 0.07 | 0.919 | 0.002 | 0.8298 |
| Cluster 5 | 12 | (-17, -89, -1) | 0.42 ± 0.06 | 0.35 ± 0.06 | 1.043 | 0.000 | 0.8189 |
| Cluster 6 | 11 | (-20, -91, 0) | 0.44 ± 0.06 | 0.38 ± 0.07 | 1.006 | 0.001 | 0.7685 |
| Cluster 7 | 48 | (30, -71, 9) | 0.59 ± 0.06 | 0.54 ± 0.05 | 0.933 | 0.002 | 0.8534 |
| **Chronic Low Back Pain > Controls** |  |  |  |  |  |  |  |
| Cluster A | 16 | (-13, 34, 38) | 0.44 ± 0.04 | 0.48 ± 0.06 | -0.818 | 0.004 | 0.8066 |
| Cluster B | 16 | (-15, 26, 42) | 0.51 ± 0.04 | 0.55 ± 0.06 | -0.857 | 0.003 | 0.8128 |
| Cluster C | 10 | (-37, 27, -8) | 0.25 ± 0.08 | 0.3 ± 0.05 | -0.885 | 0.003 | 0.8288 |
| Cluster D | 16 | (43, -48, 2) | 0.51 ± 0.04 | 0.56 ± 0.06 | -0.982 | 0.001 | 0.7816 |

<sup>a</sup>Cohen's D calculated as control FA mean minus CLBP FA mean divided by pooled CLBP and HC standard deviation (i.e. negative value indicates CLBP > control mean)
<sup>b</sup>Group differences assessed with a two sample t-test at $p < 0.05$ , uncorrected for multiple comparisons

### 3.2 Stability of Findings Across Visits

To confirm the stability of these findings, the group differences within the 11 identified clusters were examined at each visit independently. The mean FA values are shown for all 11 clusters for each subject and visit in Figure 3. The mean FA values for individual subjects remained consistent across the three visits, with minimal intra-subject variance for both control and CLBP participants.

**Figure 3.**
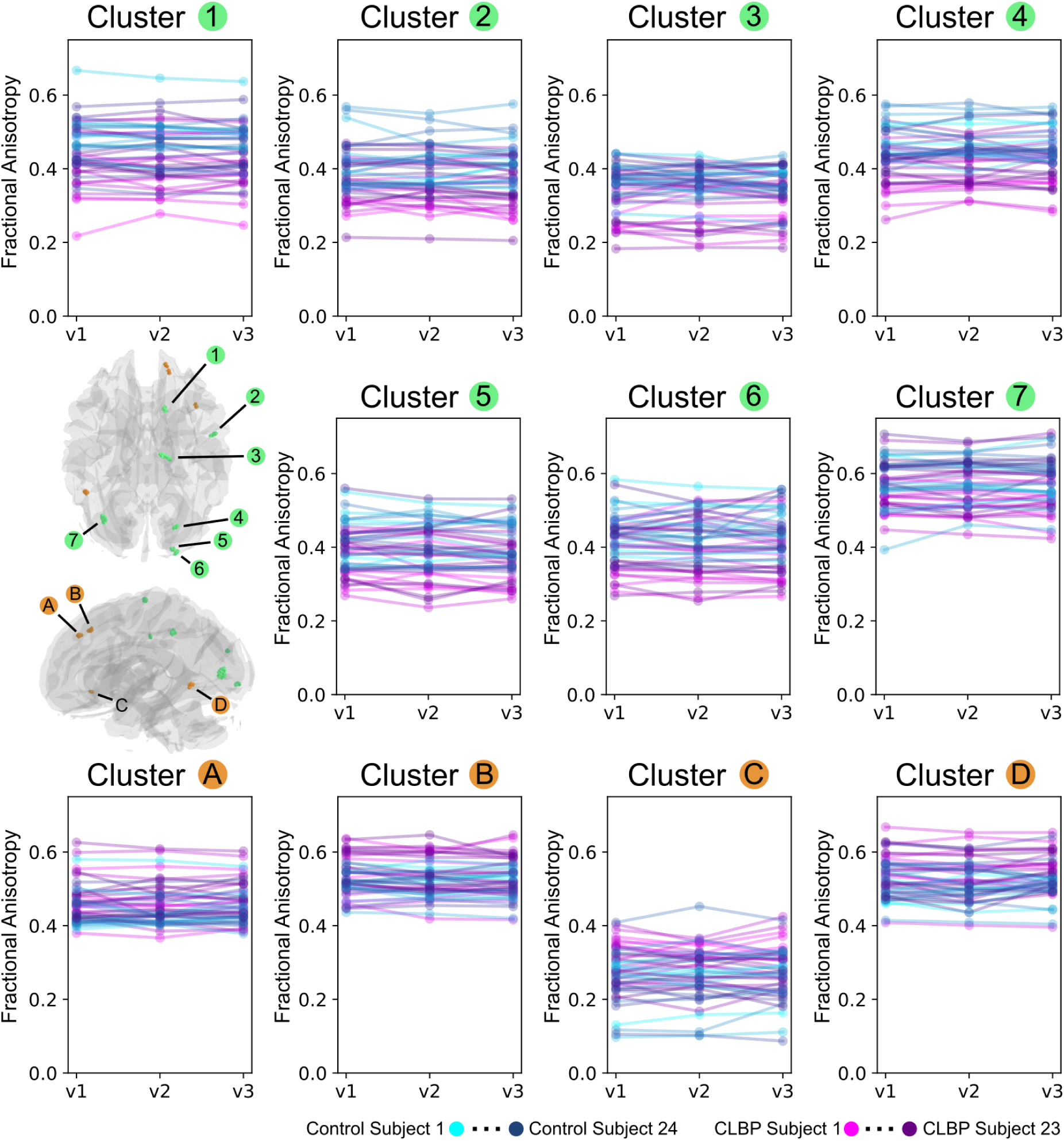
Mean fractional anisotropy calculated for regions remaining after the proposed repeatability analysis. The 7 regions where controls had higher FA relative to CLBP are shown in green and regions where CLBP had higher FA relative controls are shown in orange. Average fractional anisotropy is displayed for each visit for the 24 control subjects (separate colors cyan to navy) and 23 chronic low back pain subjects (separate colors pink to indigo) that had imaging data collected at all three visits. Values and group differences were stable across the three visits for all regions.

Table 2 provides a quantitative analysis of the stability of fractional anisotropy group differences, reporting the mean FA, Cohen’s D, and p-values for each cluster at each of the three visits. As intended by the design of the repeatability analysis, the direction of the group difference (e.g., Controls > CLBP) and the statistical significance were highly consistent for all 11 clusters across each time point. For example, Cluster 7 (FA Controls > FA CLBP) had p-values of 0.002, 0.003, and 0.004 (uncorrected for multiple comparisons) for visits 1, 2, and 3, respectively along with high I2C2 (Table 1. I2C2: 0.8534) which was used for thresholding clusters. This demonstrates that the repeatability pipeline is correctly identifying regions with consistent group differences at each visit along with the previously stated minimal intra-subject variance.

**Table 2.** Group average fractional anisotropy reported separately per visit.

| | | **Control**<br>Mean FA ± Std | **CLBP**<br>Mean FA ± Std | **Cohen's D<sup>a</sup>** | **P-Value<sup>b</sup>** |
| --- | --- | --- | --- | --- | --- |
| **Visit 1 (Control N=24, CLBP N=27)** |  |  |  |  |  |
| Control ><br>CLBP | Cluster 1 | 0.48 ± 0.07 | 0.41 ± 0.07 | 1.022 | > 0.001 |
| | Cluster 2 | 0.42 ± 0.07 | 0.36 ± 0.07 | 0.830 | 0.004 |
| | Cluster 3 | 0.37 ± 0.05 | 0.32 ± 0.07 | 0.908 | 0.002 |
| | Cluster 4 | 0.47 ± 0.05 | 0.41 ± 0.07 | 0.962 | 0.001 |
| | Cluster 5 | 0.43 ± 0.07 | 0.36 ± 0.06 | 1.076 | > 0.001 |
| | Cluster 6 | 0.45 ± 0.06 | 0.38 ± 0.06 | 1.120 | > 0.001 |
| | Cluster 7 | 0.59 ± 0.07 | 0.54 ± 0.05 | 0.914 | 0.002 |
| CLBP ><br>Control | Cluster A | 0.44 ± 0.04 | 0.48 ± 0.06 | -0.768 | 0.008 |
| | Cluster B | 0.51 ± 0.04 | 0.56 ± 0.06 | -0.841 | 0.004 |
| | Cluster C | 0.24 ± 0.08 | 0.3 ± 0.05 | -0.914 | 0.003 |
| | Cluster D | 0.51 ± 0.05 | 0.56 ± 0.05 | -1.028 | > 0.001 |
| **Visit 2 (Control N=25, CLBP N=24)** |  |  |  |  |  |
| Control ><br>CLBP | Cluster 1 | 0.47 ± 0.07 | 0.41 ± 0.07 | 0.869 | 0.004 |
| | Cluster 2 | 0.42 ± 0.06 | 0.35 ± 0.06 | 1.183 | > 0.001 |
| | Cluster 3 | 0.37 ± 0.05 | 0.32 ± 0.07 | 0.807 | 0.008 |
| | Cluster 4 | 0.47 ± 0.05 | 0.4 ± 0.05 | 1.174 | > 0.001 |
| | Cluster 5 | 0.42 ± 0.06 | 0.36 ± 0.07 | 0.968 | 0.001 |
| | Cluster 6 | 0.44 ± 0.06 | 0.38 ± 0.08 | 0.778 | 0.01 |
| | Cluster 7 | 0.59 ± 0.06 | 0.54 ± 0.06 | 0.905 | 0.003 |
| CLBP ><br>Control | Cluster A | 0.44 ± 0.04 | 0.47 ± 0.06 | -0.733 | 0.015 |
| | Cluster B | 0.51 ± 0.04 | 0.55 ± 0.06 | -0.788 | 0.009 |
| | Cluster C | 0.25 ± 0.08 | 0.3 ± 0.05 | -0.706 | 0.017 |
| | Cluster D | 0.5 ± 0.05 | 0.55 ± 0.06 | -0.803 | 0.008 |
| **Visit 3 (Control N=25, CLBP N=23)** |  |  |  |  |  |
| Control ><br>CLBP | Cluster 1 | 0.47 ± 0.07 | 0.41 ± 0.07 | 0.921 | 0.003 |
| | Cluster 2 | 0.41 ± 0.06 | 0.34 ± 0.07 | 1.182 | > 0.001 |
| | Cluster 3 | 0.36 ± 0.05 | 0.31 ± 0.07 | 0.876 | 0.005 |
| | Cluster 4 | 0.47 ± 0.05 | 0.4 ± 0.06 | 1.103 | > 0.001 |
| | Cluster 5 | 0.41 ± 0.06 | 0.35 ± 0.06 | 0.981 | 0.001 |
| | Cluster 6 | 0.45 ± 0.07 | 0.37 ± 0.07 | 1.043 | > 0.001 |
| | Cluster 7 | 0.59 ± 0.06 | 0.54 ± 0.06 | 0.869 | 0.004 |
| CLBP ><br>Control | Cluster A | 0.44 ± 0.05 | 0.48 ± 0.06 | -0.724 | 0.017 |
| | Cluster B | 0.51 ± 0.04 | 0.54 ± 0.06 | -0.727 | 0.017 |
| | Cluster C | 0.25 ± 0.07 | 0.3 ± 0.06 | -0.799 | 0.008 |
| | Cluster D | 0.51 ± 0.04 | 0.55 ± 0.06 | -0.841 | 0.006 |

<sup>a</sup>Cohen's D calculated as control FA mean minus CLBP FA mean divided by pooled CLBP and HC standard deviation (i.e. negative value indicates CLBP > control mean)
<sup>b</sup>Group differences assessed with a two sample t-test at $p < 0.05$ , uncorrected for multiple comparisons

### 3.3 Reduced Chance Findings Relative to a Single-Visit Design

A key advantage of the proposed repeatability analysis is its ability to filter out spurious or non-repeatable findings that may arise in standard cross-sectional studies. As illustrated in Figure 4, analyzing each visit separately identified a large number of significant clusters, the majority of which were not consistent across visits. Figure 4A shows an example of a repeatable cluster, where the significant regions from each visit substantially overlap with the final cluster identified by the repeatability analysis. In contrast, Figure 4B provides an example of unreliable findings, where clusters were identified at single visits but showed no spatial agreement across time.

**Figure 4.**
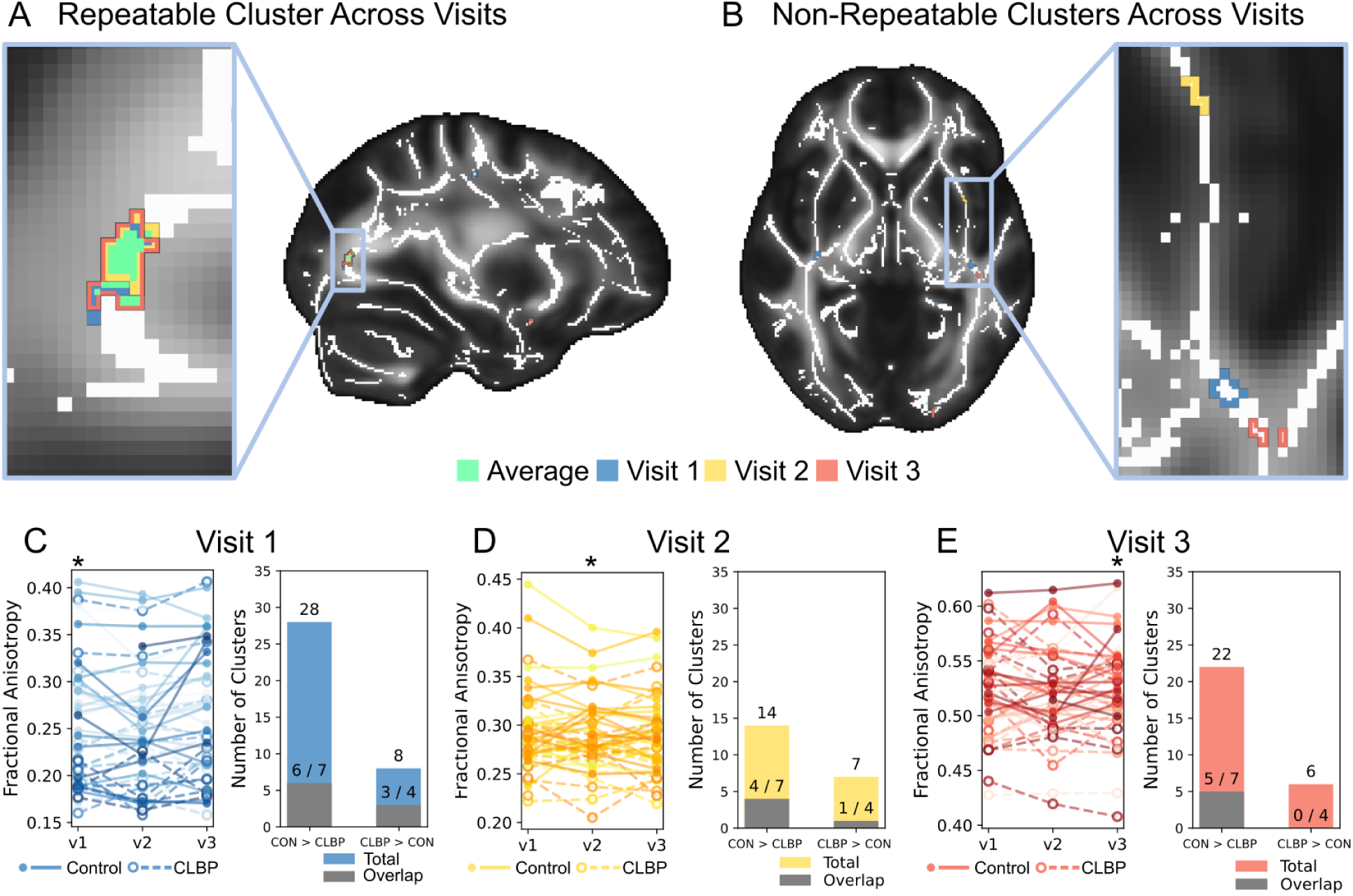
A) An example region where fractional anisotropy was observed to be higher in controls compared to chronic low back pain subjects at all visits. The cluster extracted with the proposed repeatability analysis is shown in green with overlapping clusters for visit 1 (blue), visit 2 (yellow), and visit 3 (red) overlaid. B) Shows an example of non-repeatable regions where fractional anisotropy differences were observed using t-statistics at single visits but there was no spatial agreement among visits. For each non-repeatable region in B, mean fractional anisotropy is plotted for each subject per visit (C left, D left, E left). Bar charts indicating the number of clusters detected across the entire brain for each visit (C right, D right, E right) are shown alongside the total number of clusters at each visit overlapping the 11 clusters output by the proposed repeatability analysis (grey, 7 control FA greater than CLBP FA, 4 CLBP FA greater than control FA). Although there are numerous clusters above threshold (voxelsize greater than 10, t-statistic > |2.5|) using each visit separately, the majority of these clusters are chance findings that are not repeatable across visits.

The bar charts in Figures 4C, 4D, and 4E quantify this effect. At Visit 1, a standard analysis yielded 36 clusters (28 for Controls > CLBP, 8 for CLBP > Controls), but only 9 of these (6/7 and 3/4) overlapped with the 11 final repeatable clusters. Similarly, at Visit 2, 21 clusters were found, with only 5 overlapping the final results. At Visit 3, 28 clusters were found, with only 5 overlapping. These results strongly suggest that the majority of clusters identified in a single-visit analysis are chance findings that are not stable over time, highlighting the value of the proposed repeatability-based filtering approach. Further examination revealed that clusters identified in the proposed repeatability analysis but absent in individual-visit analyses did not exceed the minimum voxel size threshold when assessed at single visits.

### 3.4 Anatomical Localization of Repeatable Findings

To place the identified clusters into anatomical context, a select subset of clusters were overlaid on major white matter tracts, as shown in Figure 5. The 4 clusters residing in the occipital lobe with higher FA in controls were situated near known sensory and visual pathways. Specifically, clusters 4, 5, and 6 are adjacent to the left optic radiation and the occipital portion of the corpus callosum, while cluster 7 is near the right optic radiation. The two selected clusters with higher FA in the CLBP group were located near frontal tracts. As shown in Figure 5B, clusters A and B are adjacent to the left fronto-pontine tract, the frontal portion of the corpus callosum, and the left frontal aslant tract. It is important to note, however, that the proximity of these clusters to regions of crossing fibers impedes their definitive localization to a single white matter tract.

**Figure 5.**
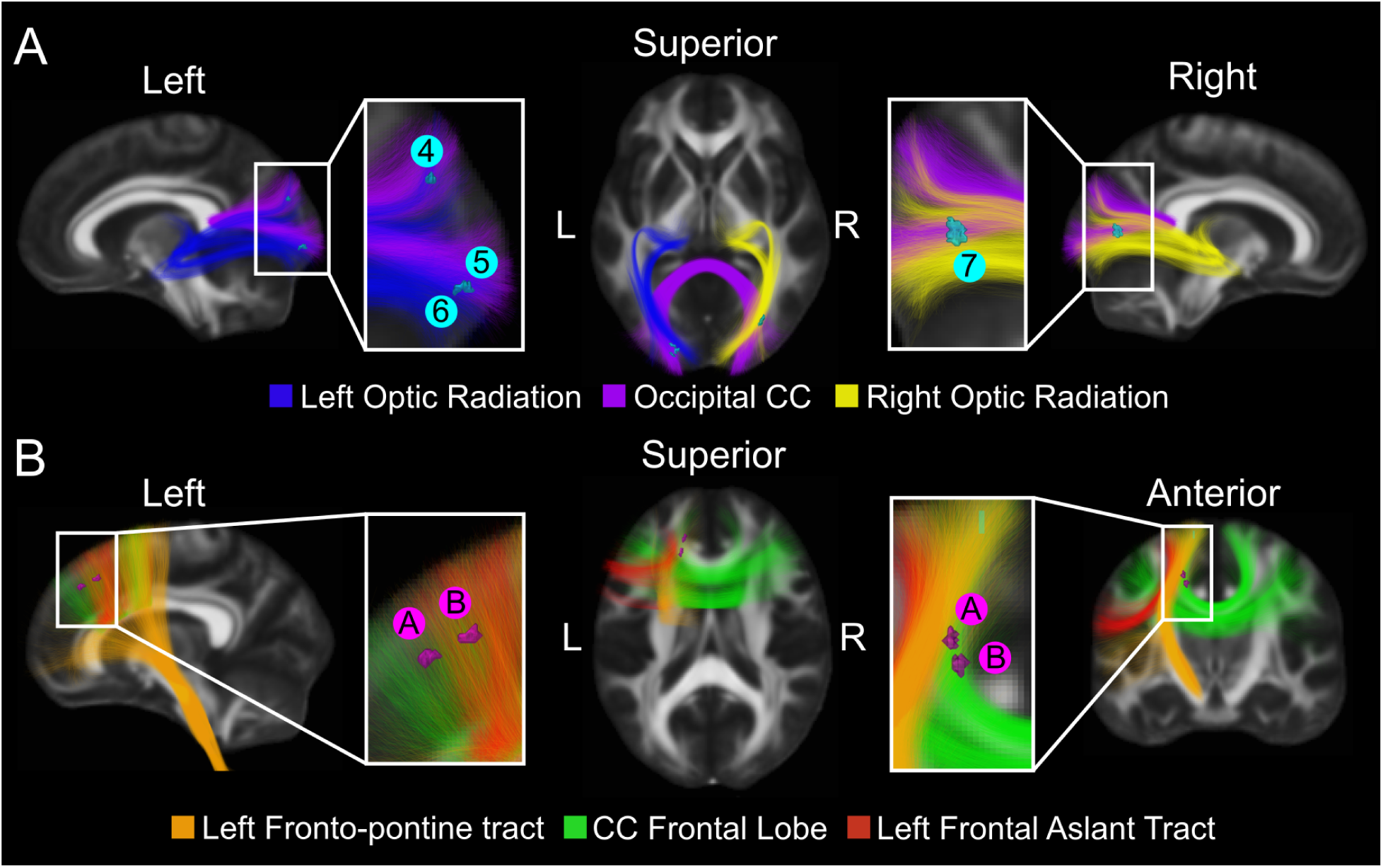
Visualization of select regions output from the repeatability analysis where a fractional anisotropy group difference was observed. A) 4 clusters where control fractional anisotropy was higher than chronic low back pain fractional anisotropy are displayed (cyan: 4, 5, 6, 7) and overlaid on adjacent white matter tracts (blue: left optic radiation; purple: occipital corpus callosum; yellow: right optic radiation). B) 2 clusters where chronic low back pain fractional anisotropy was greater than control fractional anisotropy are displayed (pink: A, B) and overlaid on adjacent white matter tracts (orange: left fronto-pontine tract; green: corpus callosum frontal posterior portion; red; left frontal aslant tract). Although regions overlap major white matter tracts, their proximity in fiber crossing regions impedes the exact localization of these regions to individual white matter tracts.

### 3.4 Comparison of Proposed Method with Alternative Tractometry Methodology

Six white matter tracts overlapped the six selected clusters from the primary analysis (four where control FA > CLBP FA and two where CLBP FA > control FA). Tract profiles for each of these six tracts are visualized per group averaged for each visit in Figure 6. Of the six tracts investigated, only the Left Optic Radiation had segments that showed statistically significant differences in fractional anisotropy (FA) between the control and CLBP groups across all three visits (p<0.05, uncorrected). While other tracts displayed segments with significant group differences, these findings were less consistent across the three time points. In regions where statistically significant differences were observed for two or less visits, all three visits still had mean differences between groups larger than standard deviations (See zoomed in regions for the right optic radiation Figure E.1 and E.2) reflecting the small statistical power related to the small sample size but consistent trend level group differences across all visits. There was a strong spatial correspondence between the two methodologies in regions where the TBSS repeatability analysis yielded a statistically different group difference (highlighted 7 light red regions Figure 6). Among the tracts investigated, 7 regions overlapped retained TBSS repeatability analysis clusters, of which 5 had adjacent segments with statistically significant tractometry group differences.

**Figure 6.**
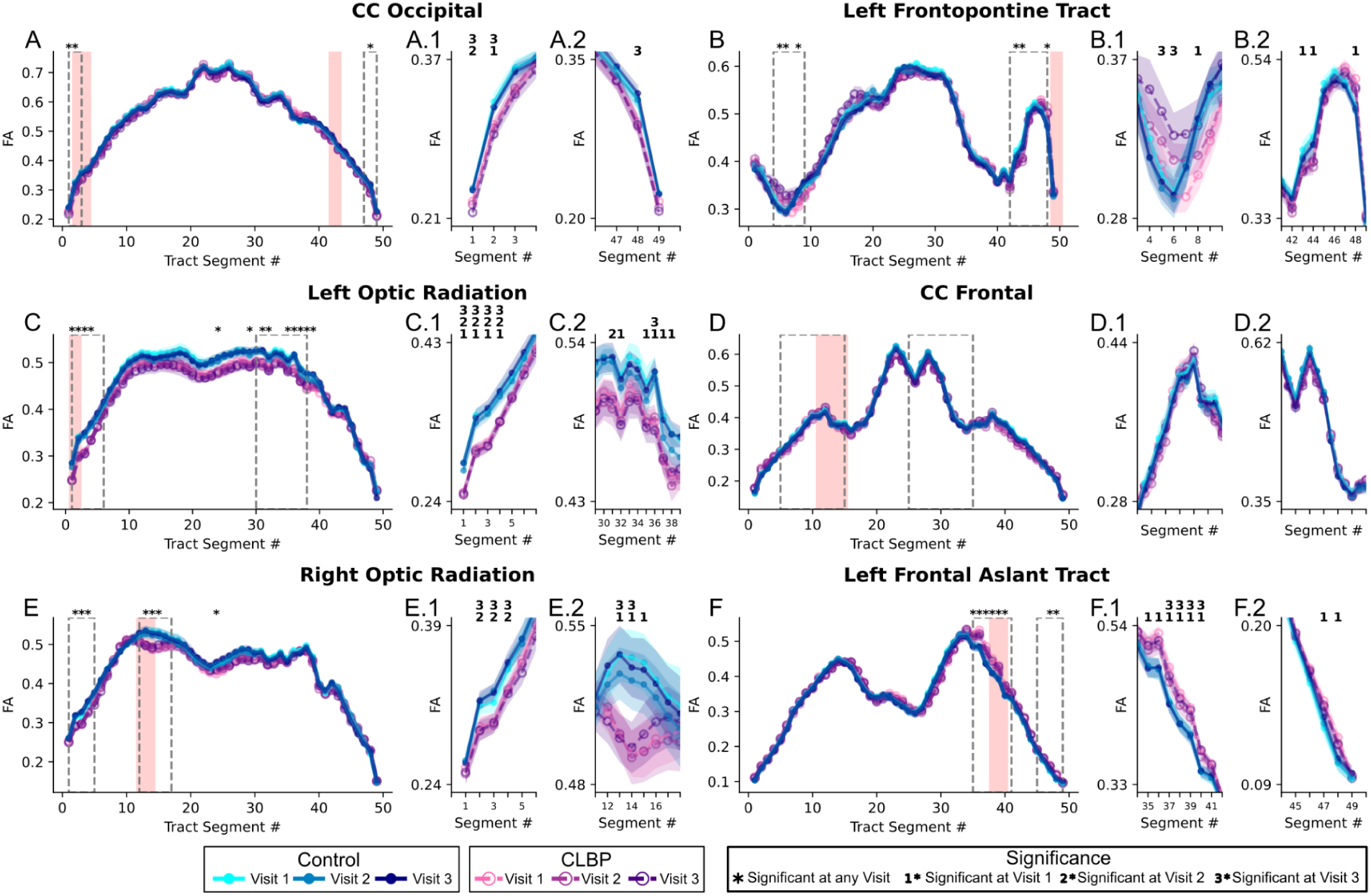
Tractometry profiles of fractional anisotropy for six selected white matter tracts that overlapped regions output from the TBSS repeatability analysis; A) Occipital Portion of the Corpus Callosum, B) Left Frontopontine Tract, C) Left Optic Radiation, D) Frontal Portion of the Corpus Callosum E) Right Optic Radiation, F) Left Frontal Aslant Tract. Average fractional anisotropy values are shown for 50 tract segments for controls (cyan to dark blue) and CLBP (pink to dark purple) with zoomed regions depicted in subplots (e.g. dotted grey boxes in A, zoomed regions in A.1, A.2). For each group and visit standard deviations are displayed with transparency in the associated color. Segments that overlap clusters identified in the proposed TBSS repeatability analysis are highlighted (light red). Regions with a statistically significant difference (two sample t-test, p < 0.05, uncorrected) for at least one of three visits are identified (*), and in the zoomed plots the specific visit with a group difference is identified with a number indicating which visit showed the significant difference.

## 4 Discussion

### 4.1 Consistent Regional Alterations in White Matter DTI metrics in Chronic Low Back Pain

The lower FA associated with CLBP and observed in 7 regions in the current study is in agreement with previous dMRI studies reporting similarly lower FA in CLBP relative to controls (Kim et al., 2020; Ma et al., 2020; Mansour et al., 2013). Four regions were found to have higher FA in CLBP relative to controls; higher FA may also be associated with CLBP. In specific contexts, lower FA may indicate demyelination as in multiple sclerosis (Inglese & Bester, 2010) but altered FA values and dMRI derived metrics should be interpreted in the context of specific diseases and could have many underlying microstructural explanations (Horsfield & Jones, 2002). Thus, given the current state of the neuroimaging CP field, it is unclear the specific microstructural explanations for such alterations of FA in CLBP. Findings from one predictive study has suggested that lower FA may indicate a transition from an acute to a chronic pain condition, suggesting the alterations in WM may exist prior to the onset of chronic pain symptomatology and with these differences measurable up to at least one year later (Mansour et al., 2013). Here, the WM matter microstructural alterations associated with CLBP were measurable at all three visits spanning 4 months, with patients presenting pain symptoms for at least 4 months prior to their first visit, supporting the finding that microstructural changes to WM are present across a long time frame after the chronifcation of pain symptoms.

### 4.2 Anatomical Localization of White Matter Alterations in CLBP

Repeatable decreases in FA associated with CLBP were found primarily in the occipital lobe (4 of 7 clusters) which has not been typically reported in CLBP but found in other chronic pain conditions such as in cluster headaches (Szabó et al., 2013; Teepker et al., 2012). The two lower FA clusters found in the current study in the superior parietal regions were located adjacent to motor regions agreeing with lower FA found in the primary somatosensory cortex white matter (Kim et al., 2020) and supports the idea that alterations to primary motor networks in these conditions are possible. While no differences were found within the internal and external capsule white matter regions as in other studies (Čeko et al., 2015; Mansour et al., 2013), higher and lower FA associated with CLBP were found in multiple regions along tracts that traverse these regions.

Isolating voxel-wise differences to singular white matter tracts was difficult because regional differences were found in areas adjacent to the cortex where multiple tracts traverse. For example, in Figure 5, the regions with lower FA in CLBP were located along the corpus callosum and left/right optic radiations, demonstrating the difficulty in interpreting these regional differences as microstructural alterations along specific tracts, as others have reported when using TBSS analysis (Ma et al., 2020; Mansour et al., 2013). Importantly, the regions with altered FA reported from TBSS analysis in the current study agreed substantially with tract segments found with altered FA extracted from tractometry, albeit often without agreement between all 3 visits. This is an encouraging finding given the differences between how FA is measured between the two methods; TBSS projects the maximum FA value from adjacent voxels to the skeleton; tractometry averages across all voxel values overlapping a segment. A likely explanation for the small discrepancies between methods is that tractometry is an average over large WM regions, whereas TBSS is a more isolated WM measurement, and highlights the importance when interpreting results extracted from differential methodology between studies.

### 4.3 Proposed Repeatability Analysis

Repeatability in the neuroimaging literature is typically reported as intra-class correlation (ICC) values and are analogous to the I2C2 values (an extension of ICC for data acquired via imaging) reported in the current study. In previous works, ICC for FA values have been reported in the good-excellent range when assessing test-retest (Cole et al., 2014; Duan et al., 2015; Rosberg et al., 2022), cross-scanner data with denoising having a positive effect on repeatability (Ades-Aron et al., 2025). This current study specifically aimed to find regions that had repeatable measurements in both controls and CLBP patients, having I2C2 values in line with previous work (I2C2 values of 0.7685 - 0.8821) in the current study. This enabled the removal of regions where CLBP vs control group differences were observed at a singular visit but not at all three time points. For example, at visit 1, 26 regions surpassed cluster size and statistics thresholds with FA values lower in CLBP patients compared to controls; however while using the proposed repeatability analysis only 7 such regions were retained. While it is notable that this repeated measures design comes with the added cost, effort and time needed to repeat the entire imaging acquisition, the technique has a substantial benefit to reduce the reporting of chance findings that can arise solely by measurement fluctuations when only single time points are considered.

### 4.4 Limitations and Future Directions

Even though the results are extremely repeatable, the study is still limited by a small sample size (25 controls, 27 CLBP) making it difficult to generalize these results to the entire CLBP population at large. This is a common limitation observed in the CLBP dMRI literature with similar sized cohorts being used previously (Čeko et al., 2015; Ma et al., 2020; Mansour et al., 2013). The small sample size also likely impeded full inter-method agreement between the TBSS and tractometry analyses; while clear, trend-level group differences were often observed across all visits in the same regions for both methods, only a subset of these differences reached statistical significance, likely reflecting low statistical power. One of the goals of the current study is to expand this pilot cohort to a larger sample size more representative of the larger population and to allow for more stringent statistical testing (e.g. cluster based statistical thresholding and multiple comparison corrections) not performed in this initial analysis.

A further limitation, inherent to the resolution of dMRI and tensor-based analysis, is the difficulty in definitively localizing observed effects to a single white matter tract, especially in regions with complex fiber crossings. Future work could address this by applying more complex modelling techniques that consider fiber crossings, such as fixel-based analysis (Dhollander et al., 2021), to disentangle the contributions of individual fiber populations within a voxel and pinpoint the specific tracts driving the observed diffusion differences.

We acknowledge that the repeated-measures design involving three separate time points is costly and difficult to implement. However, now highly repeatable clusters have been identified, a future direction could be to use these locations as a priori regions of interest. Validating these specific regions in a new, larger cohort may only require one or two scans, leveraging the knowledge gained from this longitudinal design to create more efficient and targeted studies moving forward.

## 5 Conclusion

In this study, we introduce a longitudinal dMRI study design and analysis that prioritizes the repeatability of findings to investigate WM microstructural alterations in CLBP. Here, 11 regions were identified with differences in FA between CLBP patients and healthy controls that were consistent across 3 imaging sessions spanning four months. Lower FA in CLBP was found in 7 regions whereas higher FA was found in 4 regions. Lower FA in CLBP is consistent with previous studies, although our analysis identified regions not typically reported in CLBP such as within the occipital lobe.

The primary contribution of this work is the demonstration that incorporating repeatability metrics directly into the analysis framework can substantially increase the reliability of results. Standard, single-visit analyses revealed a large number of significant clusters that were not stable across time, suggesting a high rate of chance findings. Our method effectively filters these spurious results by retaining only those regions that show both a significant group difference and high test-retest reliability. In future work, this cohort will be expanded to a larger, more representative sample, which will allow for more robust statistical testing and better generalization of these findings to the broader CLBP population. By leveraging longitudinal data to filter for repeatability, this methodology offers a path toward reliable and clinically meaningful biomarkers in chronic pain and other neurological conditions facing similar challenges of reproducibility.

## Data Availability

All data produced in the present study are available upon reasonable request to the authors

